# The impact of school closures on adolescent health-related outcomes during the COVID-19 pandemic: A natural experiment in South Korea

**DOI:** 10.1101/2021.08.12.21261943

**Authors:** Hanbin Lee, Buhm Han

## Abstract

A large number of countries implemented school closure as a response to the COVID-19 pandemic in 2020. As existing studies mostly rely on retrospective or pre/post comparisons that are prone to unmeasured confounding, the effect of school closure on adolescent health is poorly understood. The South Korean government implemented school closure to prevent the spread of COVID-19. A difference-in-differences comparing changes in health-related outcomes between provinces with differing degrees of school closure was performed. The main analysis group consists of middle school students of age 14 to 16 who were hit hardest where up to 73% of total schooling was taken online in Seoul (the physical attendance was reduced from 170 days to 45 days). For sensitivity analysis, a placebo group of high school students of age 19 who attended all school-days physically was included to detect any violation of our identification strategy. In the main analysis group of boys that experienced reduced physical school-days, both total and vigorous physical activity were reduced (−0.35 [-0.54 -0.17] days/week for vigorous physical activity and -0.38 [-0.61 -0.16] days/week for total physical activity) while such effect was absent in the placebo group of boys that actually did not experience school closure (−0.08 [-0.49 0.32] days/week for vigorous PA and -0.16 [-0.67 0.34] days/week for total PA). In girls, vigorous physical activity decreased (−0.22 [-0.40 -0.04] days/week) but the total physical activity was nearly constant (0.03 [-0.18 0.25] days/week). Other outcomes were largely unchanged.

## Introduction

The COVID-19 pandemic brought an unprecedented disruption to the public education sector in South Korea. In 2020, the physical school attendance days (physical school-days for short) of middle schools dropped as much as 135 days, from 170 days to 45 days (73% reduction). The usual spring semester that starts at March 2^nd^ was postponed until May. Even after the spring semester started, a large portion of classes were replaced by online classes.

As this disturbance persisted throughout the year, the potential harm of school closures on adolescents gained national concerns. Existing literatures emphasize the importance of public schooling in physical and psychological developments of adolescents. Thus, the harm due to the prolonged school closure could have long-term effects that might stretch into adulthood(1, 2). To prevent such harms, a study assessing the nation-wide impact of school closure is required. Yet, existing studies mostly rely on descriptive statistics or simple pre-post comparative studies, limiting the understanding of the current disaster (3-7).

In South Korea, the reduction of physical school-days varied by province, resulting a unique natural experiment. The physical school-days were the smallest in Seoul where middle school students were at school only for 45 days, while students in Jeollanam-do went to school for 133 days. By exploiting this regional variation, we measured the effect of school closure on health-related outcomes including physical activity, sleep duration, eating behavior and stress-level through a difference-in-differences strategy (8, 9). We validated our estimates by taking a placebo group, 12^th^ grade students (or high school 3^rd^ grade) who physically attended most of the school-days despite the disruption due to the pandemic, into account. In the main analysis group of boys that experienced reduced physical school-days, both total and vigorous physical activity were reduced (−0.35 [-0.54 -0.17] days/week for vigorous physical activity and -0.38 [-0.61 -0.16] days/week for total physical activity) while such effect was absent in the placebo group of boys that actually did not experience school closure (−0.08 days [-0.49 0.32] for vigorous PA and -0.16 days [-0.67 0.34] for total PA). The analysis was based on an annually collected nationally representative sample of students, the Korean Youth Risk-Behavior Web-based Survey (KYRBS)(10), providing a chance to measure the effect of school closure in a nation-wide scale.

Furthermore, we extended our analysis to evaluate the effect of physical activity reduction on body-mass index using the Wald-IV method(8, 9). We found that total and vigorous physical activity reduction did not lead to increased BMI (−0.24 [-1.14 0.67] for vigorous physical activity and -0.24 [-1.15 0.68] for total physical activity). To see whether our finding was consistent with other methods, Mendelian randomization(11) was conducted and produced similar results. Our results were consistent with a recent review of randomized controlled trials that reached a similar conclusion(12).

## Materials and Methods

### Data collection

#### Physical school-day information

The Ministry of Education of South Korea collected the school attendance days of all schools in the country through regional education offices. The data was retrieved using the national information disclosure website (http://open.go.kr). All 17 provinces provided the average physical school-days of middle schools.

#### The Korea Youth Risk-Behavior Web-based Survey (KYRBS)

The Korea Youth Risk Behavior Web-Based Survey (KYRBS) is an annually collected nation-wide survey of middle and high school students of age 14 to 19. Each year, under the administration of the Ministry of Health and the Ministry of Education, schools are randomly selected from 17 provinces. For each school, one classroom is sampled for each grade and all the students in the selected classroom attends the survey process under the supervision of the teacher. In our study, the variable used as outcomes are listed in the **Supplementary Table 1**. The data is openly available in the KYRBS website (http://www.kdca.go.kr/yhs/).

The participants from middle school (age 14-16) were the main analysis group since they experienced the school-closure the hardest. Freshmen and sophomore students in high school were excluded from the analysis since they experienced a relatively moderate level of school closure and their exact physical school-days were not disclosed by the regional education offices. The senior students in high school attended most of the school-days physically, satisfying the condition for a placebo group where the effect of school closure is likely to be zero to minimal. Finally, we used the data from 2018 to 2020 where all the data were from the same wave of data collection. Note that data from the same wave were collected using the same sampling strategy. This is summarized in **Supplementary Figure 1**.

### Statistical analysis

#### Difference-in-Differences estimation

The difference-in-differences (DiD) strategy removes time-invariant unobserved confounding by taking the difference between the outcome of two different time-points of the same unit, and then removes the unobserved time-varying confounding by taking the difference between the differences calculated in the previous step(8, 9). The classic DiD design usually exploits binary treatments such as the discrete policy change between the states. However, in our analysis, the treatment is the physical school-days, which is a continuous variable. In such cases, the estimation can be carried out by using the following regression equation:

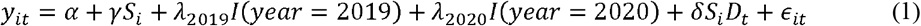

Where *y*_*it*_ is the outcome of individual *i* at time *t, S*_*i*_ is the physical school-days of individual *i* in 2020, *I(year)* are the year indicators, *D*_*t*_ is the time dummy for year 2020 and *ϵ*_*it*_ is the error term. α, γ, λ and δ are the corresponding regression coefficients.

One important assumption of the DiD strategy is the *parallel-trends assumption*. In the classic DiD design with binary treatment, this assumption requires that the counterfactual trend if not treated of the treatment group (the group in which the treatment is given) would have been the same with the observed trend of the control group. Although this assumption cannot be verified empirically, one way of evaluating the feasibility of the assumption is by looking at the pre-treatment trends (pre-trends): if the trends were to be parallel before the treatment was given, then it is likely to be parallel after the treatment. In continuous treatment DiD, the pre-trends can be assessed by estimating the following regression:

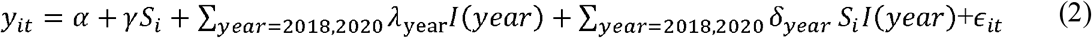

We can see if the trend from 2018 to 2019 was parallel respect to the treatment value *S*_*i*_ by checking the regression coefficient δ_2018_.

In the both regression equation (1) and (2), educational attainment (EA) of parents, the sampling region and the grade was adjusted by adding the following terms:

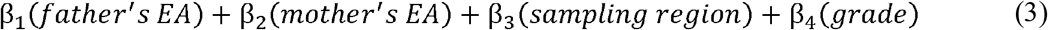

All the regression were carried out by using the R *survey*(33) package to incorporate the sampling design of the KYRBS. Note that R *survey* calculates the standard error using the sandwich estimator which is robust against heteroskedasticity and clustering.

#### Wald-IV estimation

For physical activity and body-mass index (BMI), the causal effect of school closure can be estimated through DiD. Then using the *Wald-IV* method, taking the quotient of the two-estimates, we can obtain the causal effect of physical activity on BMI as follows:

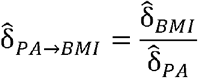

where 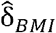 and 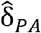 are the DiD estimates when the outcome is BMI and PA respectively. The standard error of the above estimate was calculated using the delta method. A similar approach was used in a recent study using the same dataset to evaluate the causal effect of sleep duration on BMI(14).

#### Mendelian randomization

Summary statistics of published GWAS on physical activity, childhood body mass-index and childhood obesity were used. Total two different physical activity measurements were used as exposures. Measurements from self-reported physical activity in UK Biobank were obtained from OpenGWAS(34). Two outcomes were used to measure childhood obesity. Body-mass index summary statistics came from the Norwegian Mother, Father and Child Cohort Study (MoBA)(35). Childhood obesity summary statistics came from the Early Growth Genetics Consortium(36). Genetic variants that were genome-wide significant (*P*<5×10^−8^) were selected as instruments. The details can be found in **Supplementary Table 2**. All the statistical analysis involving Mendelian randomization was performed using the TwoSampleMR(37) library.

## Results

### Characteristics of the study participants

The characteristic of the study participants can be found in **Table 1**. The average BMI was slightly higher in the placebo group than in the main analysis group. Participants in the main analysis group were more physically active than those in the placebo group. There is a large change in parental education attainment when the year changed from 2018 to 2019. This is largely due to the change in the questionnaire in 2019 where ‘do not want to respond’ became a possible option to the participants.

**Table 1.**
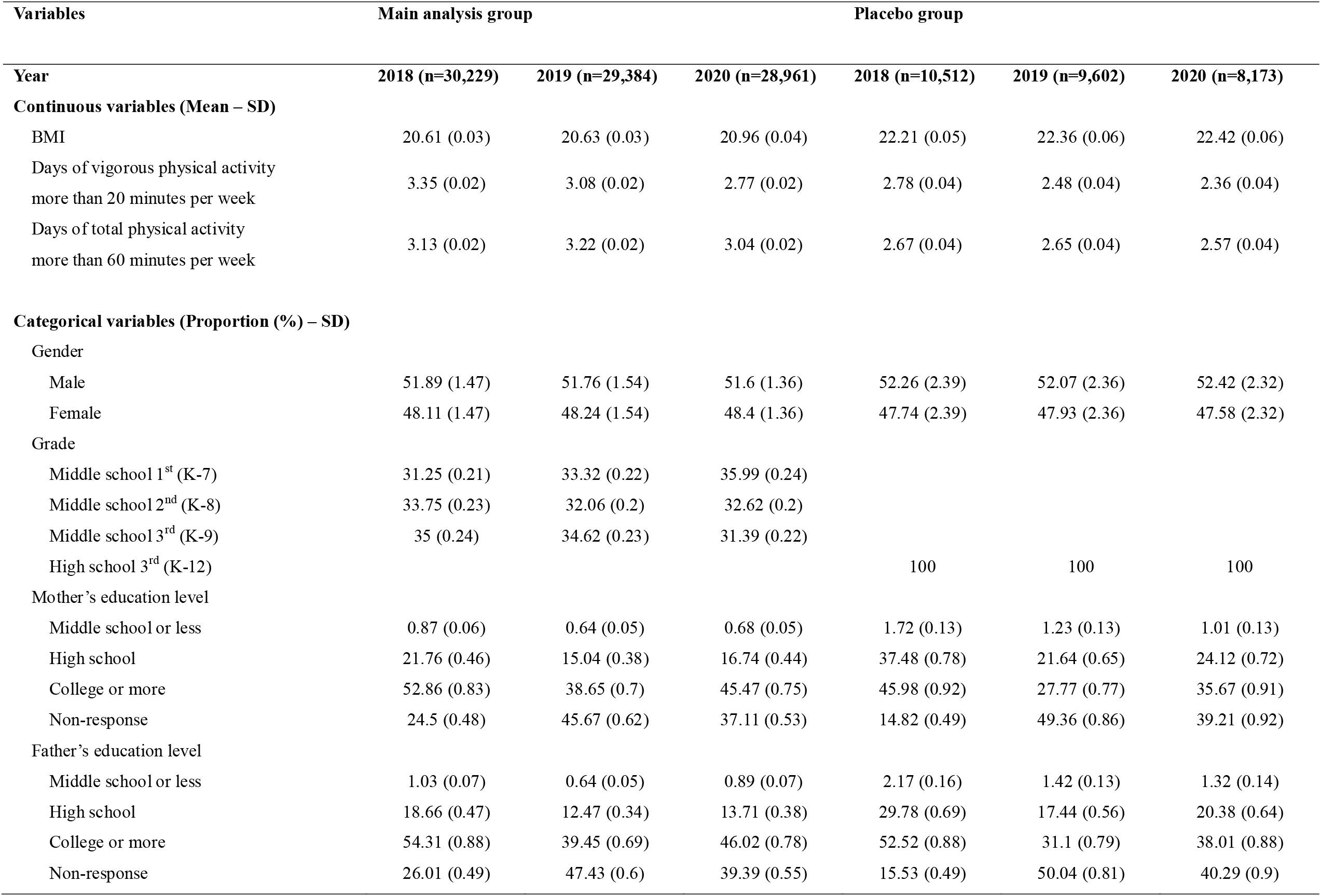
Basic characteristic of study participants

| Variables | Main analysis group |  |  | Placebo group |  |  |
| --- | --- | --- | --- | --- | --- | --- |
| Year | 2018 (n=30,229) | 2019 (n=29,384) | 2020 (n=28,961) | 2018 (n=10,512) | 2019 (n=9,602) | 2020 (n=8,173) |
| **Continuous variables (Mean – SD)** |  |  |  |  |  |  |
| BMI | 20.61 (0.03) | 20.63 (0.03) | 20.96 (0.04) | 22.21 (0.05) | 22.36 (0.06) | 22.42 (0.06) |
| Days of vigorous physical activity<br>more than 20 minutes per week | 3.35 (0.02) | 3.08 (0.02) | 2.77 (0.02) | 2.78 (0.04) | 2.48 (0.04) | 2.36 (0.04) |
| Days of total physical activity<br>more than 60 minutes per week | 3.13 (0.02) | 3.22 (0.02) | 3.04 (0.02) | 2.67 (0.04) | 2.65 (0.04) | 2.57 (0.04) |
| **Categorical variables (Proportion (%) – SD)** |  |  |  |  |  |  |
| Gender |  |  |  |  |  |  |
| Male | 51.89 (1.47) | 51.76 (1.54) | 51.6 (1.36) | 52.26 (2.39) | 52.07 (2.36) | 52.42 (2.32) |
| Female | 48.11 (1.47) | 48.24 (1.54) | 48.4 (1.36) | 47.74 (2.39) | 47.93 (2.36) | 47.58 (2.32) |
| Grade |  |  |  |  |  |  |
| Middle school 1<sup>st</sup> (K-7) | 31.25 (0.21) | 33.32 (0.22) | 35.99 (0.24) |  |  |  |
| Middle school 2<sup>nd</sup> (K-8) | 33.75 (0.23) | 32.06 (0.2) | 32.62 (0.2) |  |  |  |
| Middle school 3<sup>rd</sup> (K-9) | 35 (0.24) | 34.62 (0.23) | 31.39 (0.22) |  |  |  |
| High school 3<sup>rd</sup> (K-12) |  |  |  | 100 | 100 | 100 |
| Mother's education level |  |  |  |  |  |  |
| Middle school or less | 0.87 (0.06) | 0.64 (0.05) | 0.68 (0.05) | 1.72 (0.13) | 1.23 (0.13) | 1.01 (0.13) |
| High school | 21.76 (0.46) | 15.04 (0.38) | 16.74 (0.44) | 37.48 (0.78) | 21.64 (0.65) | 24.12 (0.72) |
| College or more | 52.86 (0.83) | 38.65 (0.7) | 45.47 (0.75) | 45.98 (0.92) | 27.77 (0.77) | 35.67 (0.91) |
| Non-response | 24.5 (0.48) | 45.67 (0.62) | 37.11 (0.53) | 14.82 (0.49) | 49.36 (0.86) | 39.21 (0.92) |
| Father's education level |  |  |  |  |  |  |
| Middle school or less | 1.03 (0.07) | 0.64 (0.05) | 0.89 (0.07) | 2.17 (0.16) | 1.42 (0.13) | 1.32 (0.14) |
| High school | 18.66 (0.47) | 12.47 (0.34) | 13.71 (0.38) | 29.78 (0.69) | 17.44 (0.56) | 20.38 (0.64) |
| College or more | 54.31 (0.88) | 39.45 (0.69) | 46.02 (0.78) | 52.52 (0.88) | 31.1 (0.79) | 38.01 (0.88) |
| Non-response | 26.01 (0.49) | 47.43 (0.6) | 39.39 (0.55) | 15.53 (0.49) | 50.04 (0.81) | 40.29 (0.9) |

### The natural experiment: school closure in South Korea during the 2020 pandemic

In South Korea, the spring semester usually starts at March 2^nd^, but in 2020, the actual starting of the semester was postponed until May. The first group of students who started to attend school were the senior students in high school (the K-12 students). As South Korea is famous for its fierce competition over admission into top universities, the College Scholastic Ability Test (CSAT) became the first concern of the public. Due to this large public concern that the school closure might have negative impacts in preparing for the CSAT, the Ministry of Education decided that the K-12 students will attend school days physically for the remaining year. This qualifies the K-12 students as a placebo group in which the school closure would not have taken effect.

The remaining students attended schools much less than the K-12 students. We selected middle school students (K-7 to 9 students) as the main analysis group because the physical school-days were smaller in middle schools than in high schools. Additionally, some regional educational offices refused to disclose the physical school-days by grade, hence, identifying the school-days of K-10 and 11 students was impossible. In **Figure 1a**, we colored the 17 provinces by their physical school-days of middle schools. The reduction of physical school-days were the largest in Seoul, where students went to school only for 45 days. The largest physical school-days was found in Jeollanam-do, where students went to school for 133 days in average.

**Figure 1.**
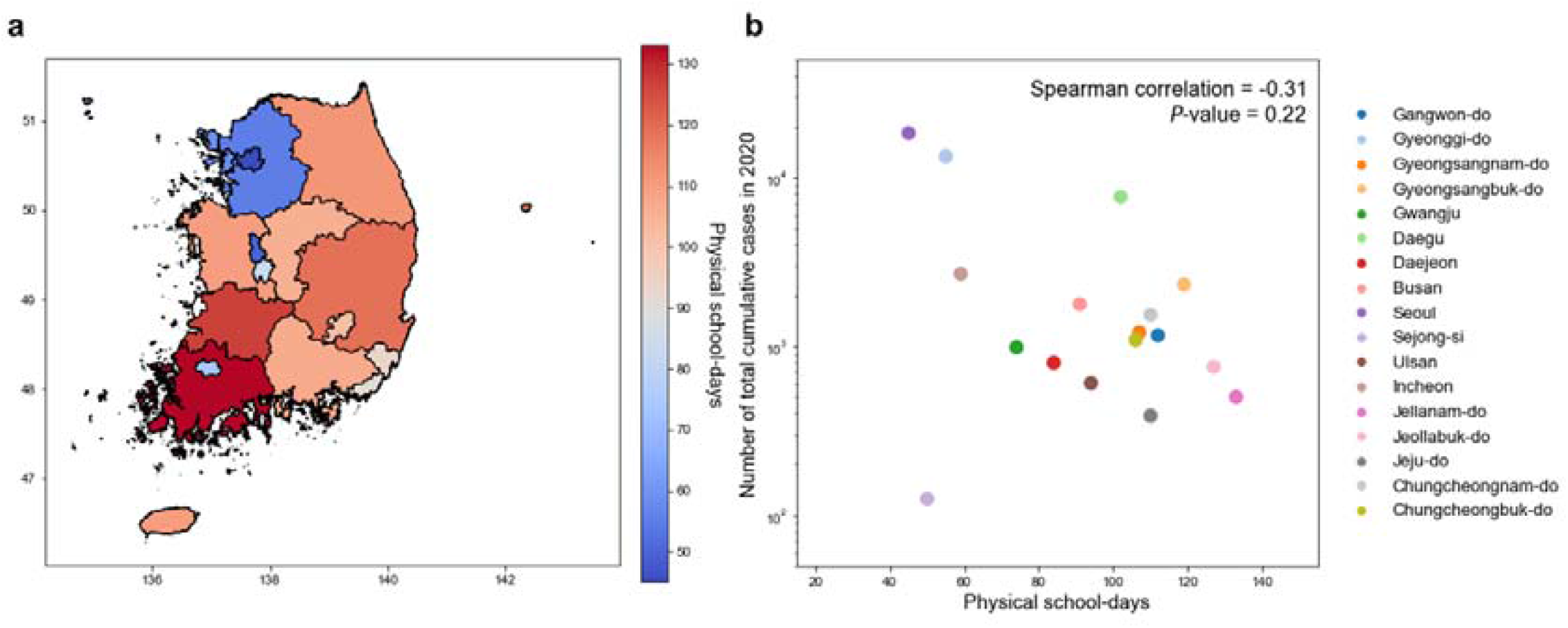
Physical school-days of 17 province in South Korea **a**. 17 provinces colored by their physical school-days. **b**. The x-axis represents the physical school-days and the y-axis represents the total cumulative number of cases in 2020.

The key assumption of the difference-in-differences (DiD) strategy is that the treatment (school closure in our study) must be exogeneous respect to the outcome(8, 9). The most likely violation of this assumption is the social distancing measures or the public fear in response to the degree of the local transmission. For example, reduced physical activity might have been the result of social distancing measures that limit outdoor activities rather than the school closure itself. Therefore, school closure must be independent from the magnitude of the local transmission in order to qualify as a proper natural experiment. We plotted the total number of cases and physical school-days to assess this possibility and found that this scenario is very unlikely (**Figure 1b**). Further analysis on cumulative cases per capita and social distancing measures showed the same result (**Supplementary Figure 2**).

### The impact of school closure in health-related outcomes

The analysis on boys and girls showed different results. The most prominent findings in boys are the large reduction in physical activity participation. The days participating in vigorous physical activity for more than 20 minutes (−0.35 days [-0.54 -0.17]) and any physical activity for more than 60 minutes (−0.38 days [-0.61 -0.16]) were significantly reduced in boys. Note that the unit is measured in change per 100 days reduction in physical school days. In girls, such reductions were weaker or absent (−0.22 days [-0.4 -0.04] for vigorous PA and 0.03 days [-0.18 0.25] for total PA). When we looked at the placebo group of boys, unlike the main analysis group of boys, these findings were absent making the findings in the main analysis group more plausible (−0.08 days [-0.49 0.32] for vigorous PA and -0.16 days [-0.67 0.34] for total PA). Drop in muscle strengthening exercise was also present in boys in the main analysis group (−0.24 [-0.43 -0.05] days/week) and was absent in the placebo group (−0.18 [-0.57 0.22]). Sedentary time dropped only in girls (−31.86 minutes [-57.77 -5.96]). However, we found a positive impact of school closure in sedentary time (68.42 minutes [9.19 127.65]) in the placebo group, making the interpretation difficult.

Eating behaviors were mostly unchanged while the sweet drink consumption has dropped in both genders (−0.18 [-0.31 -0.05] times/week and -0.28 [-0.41 -0.16] times/week respectively in boys and girls). Unlike physical activity, however, such reduction was also present in the placebo group of boys making the negative impact of school closure on sweet drink consumption likely to be a result of another event that occurred together with school closure (−0.27 [-0.52 -0.02] times/week). As not only the direction but also the effect size was similar, it is very unlikely that sweet drink dropped as a consequence of school closure. In girls, however, the placebo group exhibited significant effect in the 2018 lead which indicated a deviation in the pre-trend. Body-mass index remained stable over time as well as 5-point Likert scale stress level. Sleep duration was also unchanged.

One important feature in the DiD estimates presented in **Figure 2 and Supplementary Figure 3**, is the pre-trends from 2018 to 2019. As we have described in the **Materials and Methods** section, pre-trend can be evaluated by looking at the regression coefficient of the interaction term between the 2019 indicator and school-closure days variable. If this regression coefficient does not deviate largely from zero, we can think that the parallel trends assumption holds according to the pre-trend. In our analysis, the two physical activity outcomes have insignificant coefficients that are nearly zero, making the parallel trends assumption plausible. We see that in the main analysis group, the pre-trends are insignificant, and accordingly, the parallel trends assumption is likely to hold. On the other hand, significant pre-trends are present in the placebo group of girls (Frequency of having fastfood and Frequency of having sweet drinks) making the DiD estimates unreliable in this group.

In sum, we find that the causal impact of school closure on health-related outcomes are mostly minimal except for change in physical activity. We leave the remaining interpretation of the result to the **Discussion** section.

**Figure 2.**
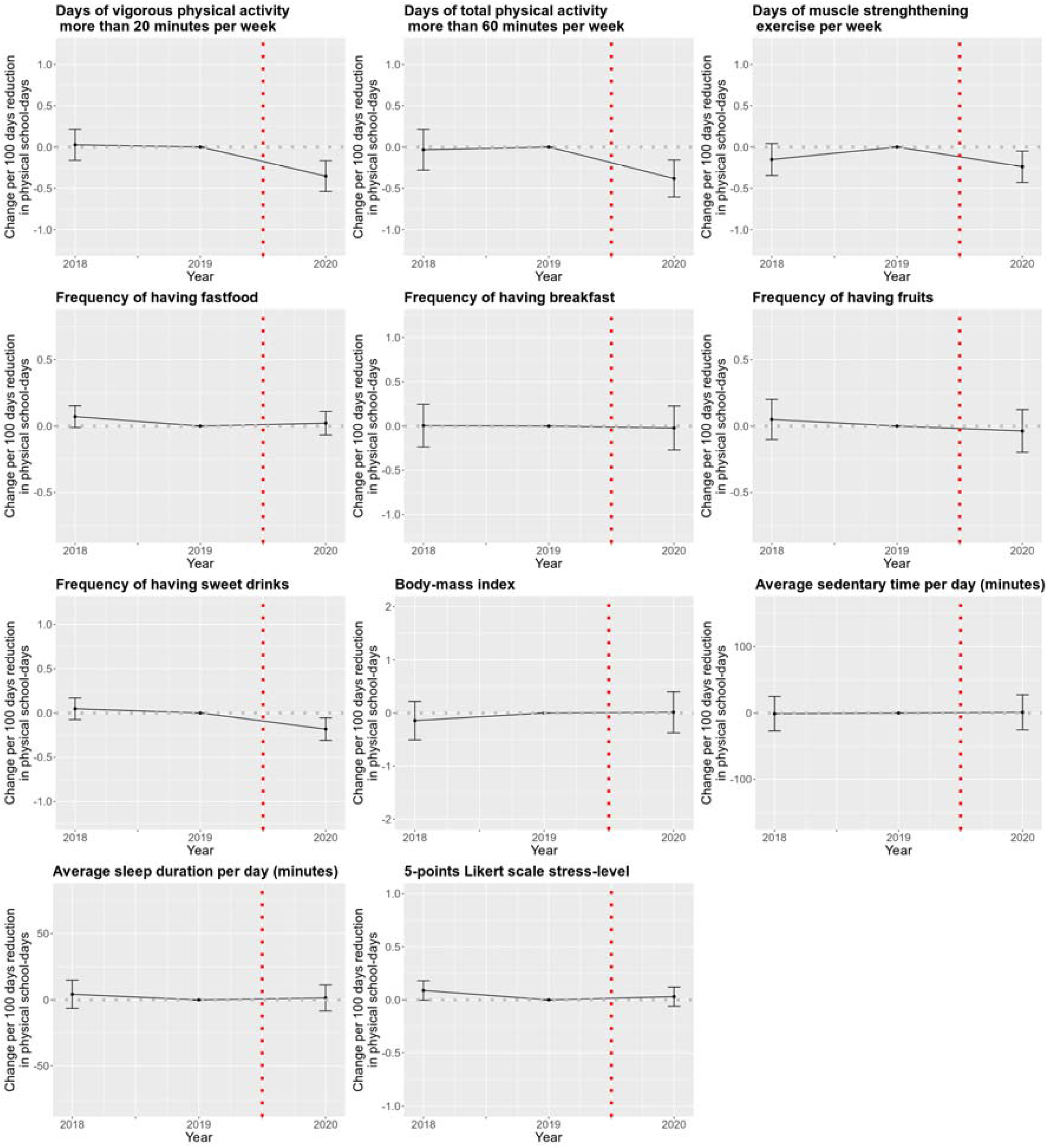
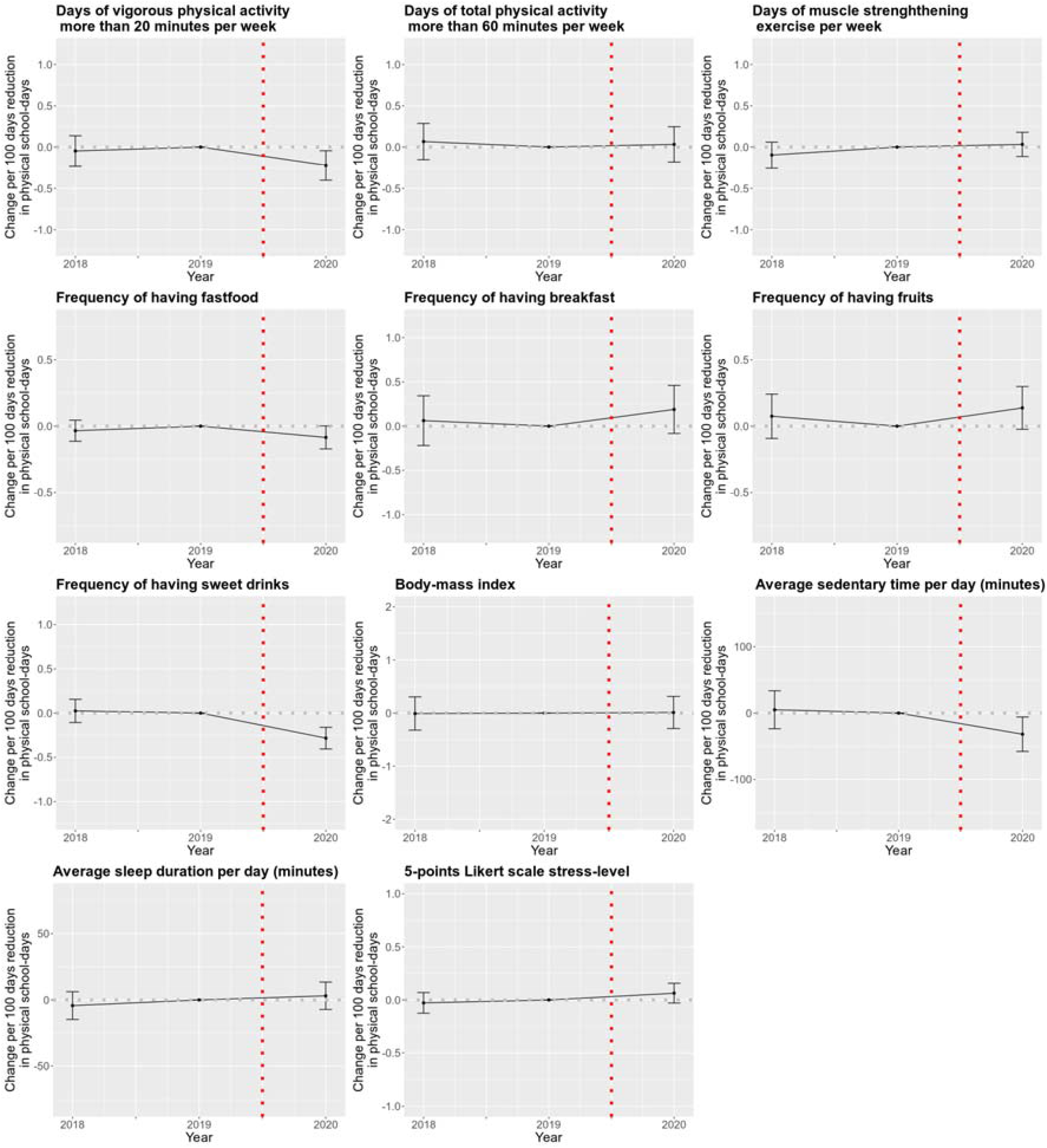
The difference-in-differences (DiD) estimates in boys (top) and girls (bottom) of the main analysis group. The bars represent the 95% confidence intervals of the regression coefficient.

### The causal effect of physical activity on body-mass index in adolescents

There is a large public effort to reduce childhood obesity in the population level, and physical activity promotion is one of the most famous means together with dietary interventions. However, contrary to common beliefs, randomized controlled trials shows minimal to no effect of increased physical activity on BMI(12). Large heterogeneity and small sample size of RCTs are other two issues making the interpretation of the study results difficult. Here, we evaluate the causal effect of physical activity on body-mass index in adolescents using school closure as an instrumental variable (IV). Our design can complement RCTs by assessing the effect of physical activity in a population level.

The three key assumptions of the IV approach are (1) relevance, (2) exogeneity and (3) the exclusion criteria (**Supplementary Figure 4**). The first assumption is met in the sample of boys. We found that the *F*-statistics were large enough to avoid the weak instrument bias (*F*-statistics =21.2 and 13.8 respectively)(13). The exogeneity assumption is impossible to empirically verify, but as we have shown in the previous chapters, likely violation due to local transmission was excluded. Additionally, time-invariant unobserved confounding and time-variant unobserved confounding obeying the parallel-trends assumption are removed since we used DiD to estimate the effect of the instrument on the outcome. The last assumption, the exclusion criteria can be violated when school closure has a direct effect on BMI independent of physical activity. Eating behavior is an obvious candidate for such violation, and sleep duration and stress level were also known to cause change in BMI(14, 15). Nevertheless, as these variables were unchanged due to school closure as in our DiD estimates, the likelihood of these variables as a violation of the exclusion criteria is very small.

An overlooked assumption when applying IV is the monotonicity assumption(16, 17). The monotonicity assumption is crucial in identifying the local average treatment effect (LATE) which is the average treatment effect in compilers (participants that conform to the change in IV). The assumption requires that there are no defiers in the sample, which means in our case, no one among the sample participated in more physical activity as a result of physical school-day reduction. Although this assumption cannot be tested directly in our sample, we argue that this assumption is likely to hold according to the schooling policy during the 2020 pandemic.

Although students did not attend schools physically, the classes were replaced by online classes. Class attendance was checked by the teachers, and hence students were forced to take classes even though they were not physically present at the school. As more than 3 hours of physical activity and exercise classes are included in the middle and high school syllabus, transition to online classes replaced these physical activities to relatively inactive classes by forcing students to stay in their houses (especially in front of their digital devices). Because middle school classes end after 4PM, and due to number of social factors in South Korea (prevalent private tutoring institutes, insufficient PA infrastructure and social distancing measures), after school physical activity is not likely to have fully compensated this loss in at-school physical activities. Hence, we believe that the monotonicity assumption is likely to hold in our sample.

As we have observed a large reduction in physical activity participation in boys, we use this as an instrument to estimate the effect of physical activity on BMI. The Wald-IV estimate of two physical activity exposures on BMI were -0.24 (−1.14 0.67) and -0.24 (−1.15 0.68) respectively (**Table 2**). To further validate our findings, we performed Mendelian randomization. We used two obesity related traits defined by BMI, BMI from the MoBA study measured at age 8 and obesity (defined by BMI) from the EGG consortium measured from age 2 to 18. The results from mendelian randomization were largely consistent with our findings using the school closure natural experiment. All estimates returned null results except for the MR-Egger estimate when the exposure was self-reported moderate physical activity and the outcome was obesity.

**Table 2.** Effect of physical activity on body-mass index

| **Method - Dataset** | **Estimate**<br>**(95% confidence interval)** |
| --- | --- |
| **Wald-IV estimate (KYRBS)** |  |
| Number of days of vigorous physical activity (>20 minutes per day) per week | -0.24 (-1.14 0.67) |
| Number of days of total physical activity (>60 minutes per day) per week | -0.24 (-1.15 0.68) |
| **Two-sample MR** |  |
| **UKB Vigorous PA – MoBA Body-mass Index** |  |
| Inverse variance weighted (IVW) estimator | 0.45 (-0.08 0.98) |
| MR-Egger | -1.28 (-5.43 2.86) |
| Weighted median estimator | 0.25 (-0.47 0.97) |
| Weighted mode estimator | 0.10 (-1.03 1.22) |
| **UKB Moderate PA – MoBA Body-mass Index** |  |
| Inverse variance weighted (IVW) estimator | -0.10 (-0.49 0.28) |
| MR-Egger | -0.63 (-3.27 2.01) |
| Weighted median estimator | -0.09 (-0.62 0.44) |
| Weighted mode estimator | -0.03 (-0.96 0.91) |
| **UKB Vigorous PA – EGG consortium Obesity** |  |
| Inverse variance weighted (IVW) estimator | -0.20 (-0.81 0.40) |
| MR-Egger | -3.74 (-8.34 0.87) |
| Weighted median estimator | 0.25 (-0.47 0.97) |
| Weighted mode estimator | 0.10 (-1.03 1.22) |
| **UKB Moderate PA – EGG consortium Obesity** |  |
| Inverse variance weighted (IVW) estimator | -0.02 (-0.40 0.35) |
| MR-Egger | -3.25 (-5.13 -1.37) |
| Weighted median estimator | 0.11 (-0.35 0.58) |
| Weighted mode estimator | 0.43 (-0.47 1.32) |

\* UKB Vigorous PA: Number of days/week of vigorous physical activity 10+ minutes, UKB Moderate PA: Number of days/week of moderate physical activity 10+ minutes

## Discussion

We evaluated the impact of school closure during the COVID-19 pandemic on adolescent health-related outcomes through a unique natural experiment in South Korea. As recognizing the extent of negative impacts of social distancing measures is a crucial part in future policy making, our study adds an important causal evidence to existing literatures on the side effects of school closure on adolescent health. Our findings suggest that school closure that persisted throughout the year had reduced physical activity significantly, however, other variables remain largely unchanged. Based on these findings, we additionally verified minimal to no effect of physical activity on adolescent BMI which was consistent with previous findings based on randomized controlled trials (RCTs)(12). Such consistency with previous literatures strengthens the validity of the current study.

This natural experiment was unique and valuable as most countries that were hit hard by the pandemic implemented very harsh social distancing measures such as the nation-wide lockdown that made commonly used causal inference techniques invalid. Under such circumstances, every unit is likely to receive the same level of treatment (e.g. nation-wide restaurant closures) limiting causal inference(18). Furthermore, cooccurrence of simultaneous policy changes at the same time and place will unfortunately eliminate the chance to separate the effect of interest(19). In South Korea, school closure was implemented in a more principled manner, where the school opening policy was consistent throughout the year(20). Thanks to this principled policy, we were able to identify the causal effect of school closure with a proper placebo group and a treatment that was independent with the local transmission level in each province.

Despite the growing concerns on the side effects of prolonged school closure and earlier studies supporting these concerns in South Korea(3, 21, 22), our findings suggest that school closure itself is not responsible for the changes in physical and mental fitness during the pandemic. Changes in physical activity was the only change that was found significant. As these studies rely on simple pre/post comparisons or retrospective cohort designs, they are prone to unobserved confounding(9, 17, 19). This underscores the importance of properly designed studies that are robust to such biases. Furthermore, future policy must be guided by these robust studies rather than relying on poorly designed ones that merely shows association than causation. This suggest that when targeting side effects of school closure, other outcomes such as academic achievement and learning are likely to have higher priority than health-related outcomes as suggested by a previous study in the Netherlands (23). However, subjective academic performance was largely constant in 2020 compared to 2018 and 2019 in the KYRBS dataset, possibly due to heterogenous nature of school closure across different countries or the self-reported nature of the data used (**Supplementary Table 3**).

The physical activity reduction that was more prominent in males are consistent with the previous findings that inactivity of female adolescents is extremely high in South Korea(24, 25). Existing studies and government investigations report that adherence to physical activity is low in female students during school classes than in males(26, 27). Therefore, the smaller effect of school closure found in girls are likely to be a consequence of pre-existing inactivity of girls before the pandemic started. This suggests that a proper policy to promote physical activity of girls are required regardless of the pandemic.

We also found that physical activity in general population of adolescents have no overall effect on BMI. Although this finding looks counter-intuitive at first glance, our additional analysis based on Mendelian randomization also supports our finding. This is also consistent with RCT systematic review by the Cochrane library(12) which found the effect of physical activity is minimal to null. Since trials are relatively small sized and short in follow-up time, our study adds a population-wide and a long-term evidence that physical activity is likely to be ineffective in reducing adolescent BMI.

Therefore, consistent results across multiple study design strongly supports our finding(28). However, this does not necessarily lead to the conclusion that physical activity is not important for physical fitness of adolescents. Existing trials show that although BMI itself does not change, more specific characters such as body composition and cardiovascular fitness(29). This suggests that BMI might be an insufficient measure to evaluate the physical fitness in adolescents (30-32).

Despite these achievements, our study also entails several limitations. First, our study is based on self-reported questionnaire that might entail a substantial measurement error. However, a study by maintainers of the data found that the responses of KYRBS are generally consistent with objective measurements(10). Second, causal inference methods, including the ones that we used, rely on number of empirically unverifiable assumptions. Although we carefully examined the possible violations by examining the pre-trends and the placebo groups, some violation might remain that can bias the estimates. Third, our study was limited only to few numbers of health-related outcome due to the insufficiency of the data. Further studies with more extensive set of variables are required to understand the full extent of the effect of school closure on adolescent health. Fourth, the Wald-IV approach only estimates the causal effect of physical activity among compliers of the intervention (local average treatment effect, LATE). The effect of physical activity on BMI may differ in always-takers and never-takers in our sample. Finally, the exposure instruments in mendelian randomization were measured in adults whereby the effect of SNPs might differ in adults and adolescents.

Nevertheless, we provide one of the first evidence up to our knowledge on the causal impact of school closure based on a nation-wide sample of adolescents. Further studies from different countries under different cultural and socioeconomic settings are required to fully understand the nature of the effect of school closure on adolescent health.

## Supporting information

STROBE checklist

## Data Availability

All data used in the study are publicly available at the Korea Disease Control and Prevention Agency (KDCA).

## Acknowledgement

We thank Dr. Ji-Yeob Choi for helpful feedback and comments.

Results on BMI from birth to childhood have been contributed by the Centre For Diabetes Research, University of Bergen, Norway, and the Norwegian Mother, Father and Child study, and has been downloaded from: https://www.fhi.no/en/studies/moba/for-forskere-artikler/gwas-data-from-moba/

This work was supported by SNU Student-Directed Education Undergraduate Research Program through the Faculty of Liberal Education, Seoul National University (2021).

