## Supplementary Figures and Tables for "The impact of school closures on adolescent health-related outcomes during the COVID-19 pandemic: A natural experiment in South Korea"

**Supplementary Table 1.** Description of outcome variables used in the study

| **Variables** | **Description** |
| --- | --- |
| Days of vigorous physical activity more than 20 minutes per week | Number of days in the past week participating in vigorous intensity physical activity for more than 20 minutes:  1- None/2- Once/3- Twice/4- 3 times/5- 4 times/6- 5 times/7- 6 times/8- 7 times |
| Days of total physical activity more than 60 minutes per week | Number of days in the past week participating in total physical activity for more than 60 minutes:  1- None/2- Once/3- Twice/4- 3 times/5- 4 times/6- 5 times/7- 6 times/8- 7 times |
| Days of muscle strengthening exercise per week | Number of days in the past week participating muscle strengthening physical activity:  1- None/2- Once/3- Twice/4- 3 times/5- 4 times/6- more than 5 times |
| Frequency of having fast food | Number of days eating fastfood in the past week:  1- None/2- Once-Twice/3- 3-4 times/4- 5-6 times/5- once daily/6- twice daily/7- More than 3 times daily |
| Frequency of having breakfast | Number of days eating breakfast in the pask week:  1- None/2- Once/3- Twice/4- 3 times/5- 4 times/6- 5 times/7- 6 times/8- 7 times |
| Frequency of having fruits | Number of days eating fruits in the past week:  1- None/2- Once-Twice/3- 3-4 times/4- 5-6 times/5- once daily/6- twice daily/7- More than 3 times daily |
| Frequency of having sweet drinks | Number of days drinking soft drinks in the past week:  1- None/2- Once-Twice/3- 3-4 times/4- 5-6 times/5- once daily/6- twice daily/7- More than 3 times daily |
| Body-mass index | Body weight (kg) / Height^2^ (m^2^) |
| Average sedentary time per day (minutes) | The average sedentary time during the last week |
| Average sleep duration per day (minutes) | The average sleep duration during the last week |
| 5-points Likert scale stress-level | Self-reported subjective stress level in 5-point Likert scale |

**Supplementary Table 2.** List of genetic instruments used in each Mendelian randomization analysis

| Exposure – Outcome | List of SNPs |
| --- | --- |
| UKB Vigorous PA –  MoBA Body-mass Index | rs1491872,rs2005617,rs2189464,rs2764261,  rs328900,rs382210,rs429358,rs6533635,rs6955240,rs7072776,rs7749823 |
| UKB Moderate PA –  MoBA Body-mass Index | rs10098073,rs1036800,rs11749912,rs11913445,rs2246122,rs3094622,  rs3129962,rs34555420,rs34775997,rs404907,rs4129572,rs429358,  rs4540651,rs4886868,rs7229874,rs7565480,rs9533455,rs997467 |
| UKB Vigorous PA –  EGG consortium Obesity | rs1491872,rs2005617,rs2189464,rs2764261,rs328900,rs382210,rs429358,  rs6533635,rs6955240,rs7072776,rs7749823 |
| UKB Moderate PA –  EGG consortium Obesity | rs10098073,rs1036800,rs11749912,rs11913445,rs2246122,rs3094622,  rs3129962,rs34555420,rs34775997,rs404907,rs4129572,rs429358,  rs4540651,rs4886868,rs7229874,rs7565480,rs9533455,rs997467 |

**Supplementary Table 3.** The effect of school closure on subjective academic performance

| **Gender/Year** | **2018** | **2019** | **2020** |
| --- | --- | --- | --- |
| **Boys** | 0.01 [-0.09 0.11] | (Reference) | -0.02 [-0.13 0.10] |
| **Girls** | 0.01 [-0.08 0.11] | (Reference) | 0.03 [-0.07 0.13] |

* The values in the table are the regression coefficients of the difference-in-differences estimation where the 5-point Likert scale subjective academic performance was the outcome. The same method was used as in other outcomes.

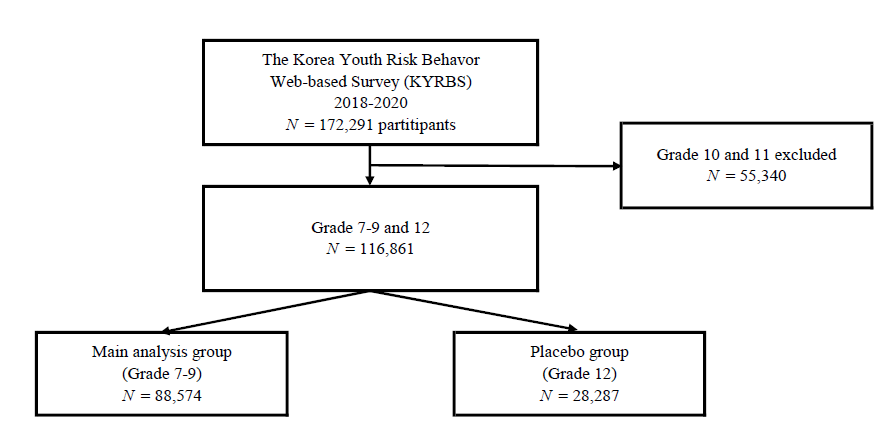

**Supplementary Figure 1.** KYRBS study participants included in the current study.

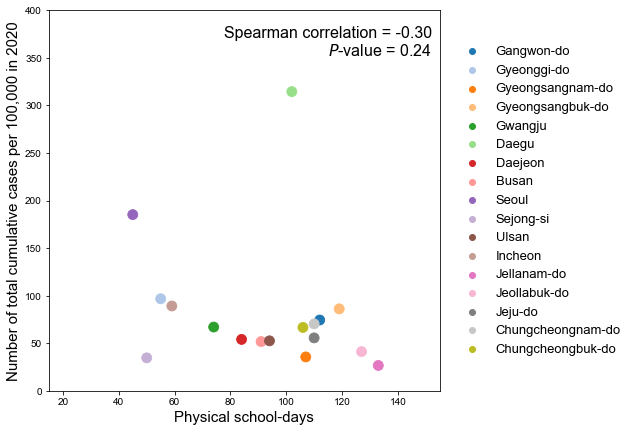

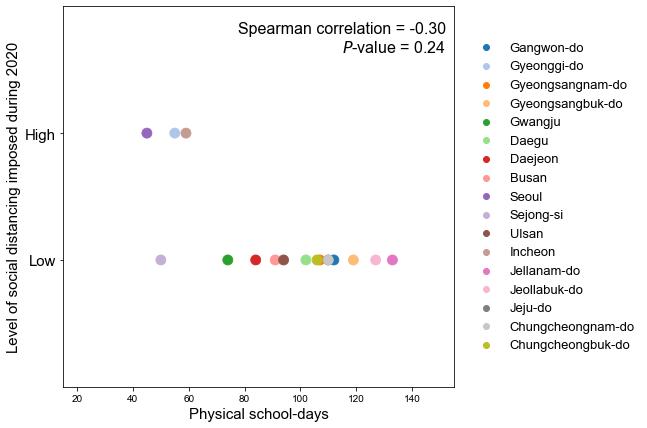

**Supplementary Figure 2.** Relationship between physical school-days and social-distancing measures during the 2020 pandemic in South Korea.

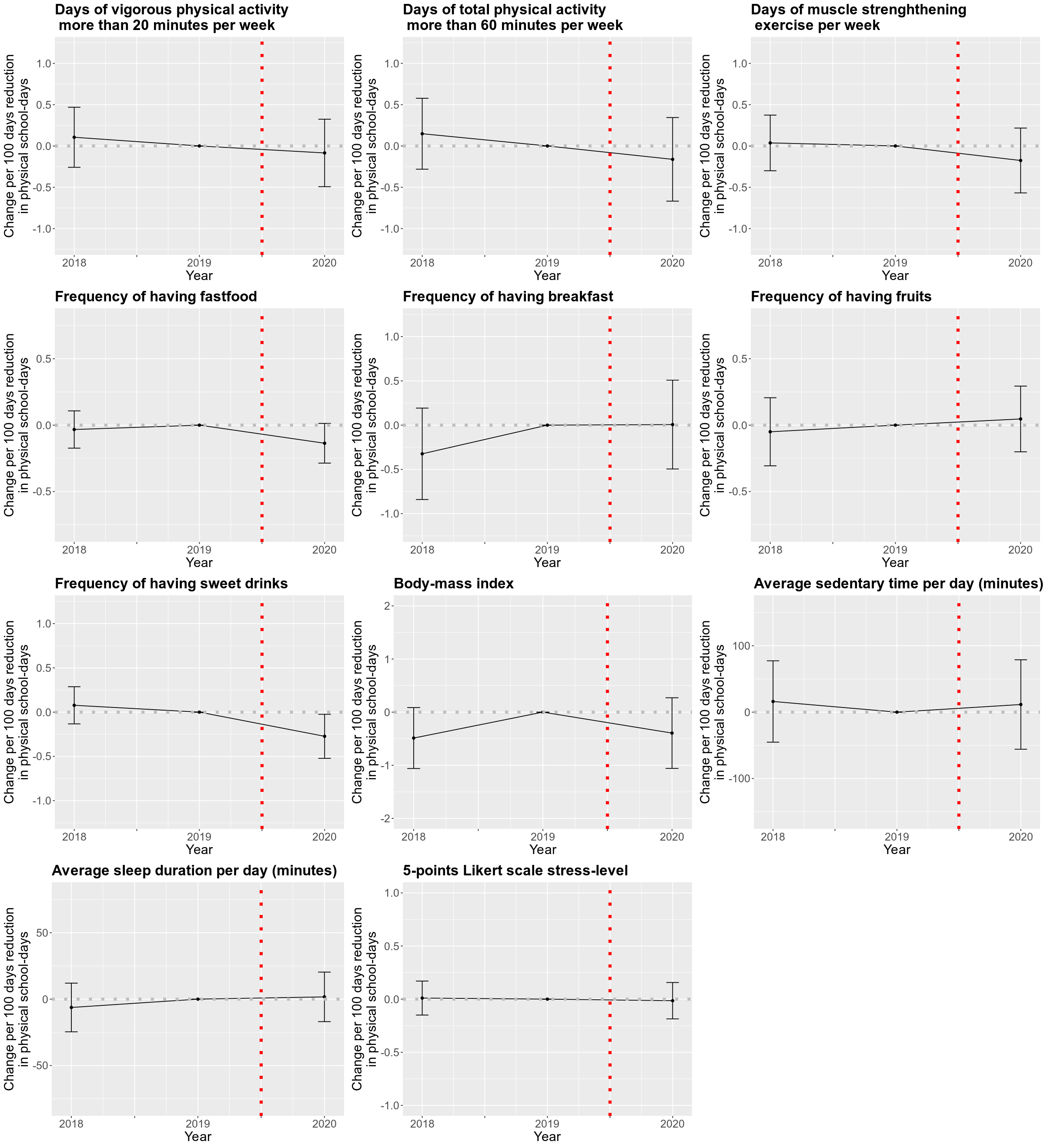

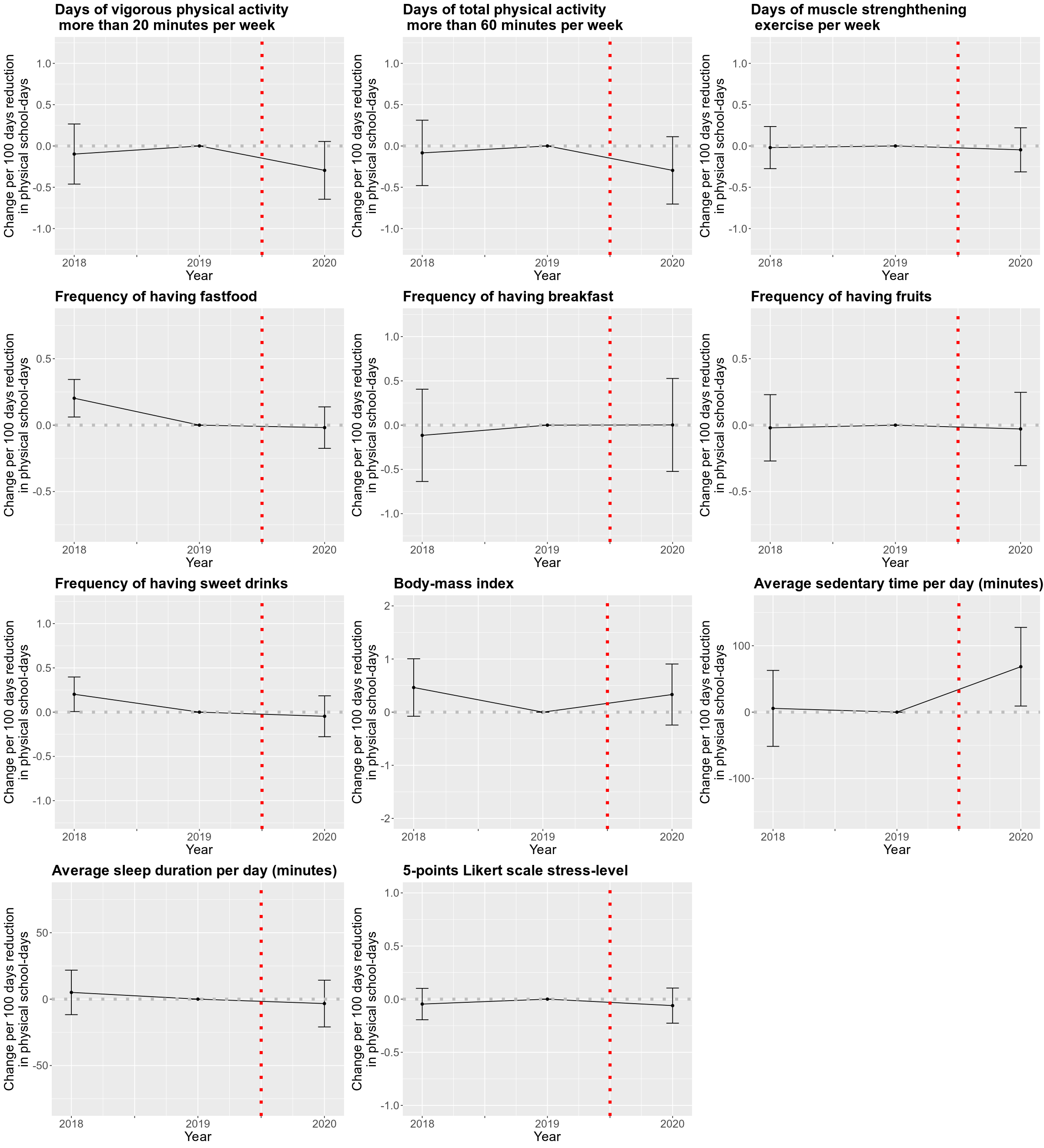

**Supplementary Figure 3.** Treatment effects of school closure on the placebo group. Boys (top) and girls (bottom).

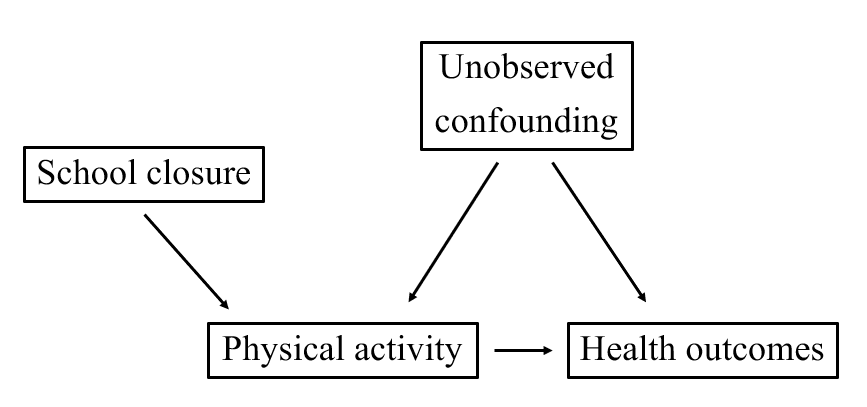

**Supplementary Figure 4.** A directed acyclic graph (DAG) depicting our instrumental variable (IV) approach.
